# Stroke Hospitalization Administration & Monitoring: Routine Or Covid-19 Care (SHAMROCC)

**DOI:** 10.1101/2023.05.29.23290701

**Authors:** Timothé Langlois-Thérien, Michel Shamy, Brian Dewar, Tim Ramsay, Ronda Lun, Dylan Blacquière, Robert Fahed, Dar Dowlatshahi, Grant Stotts, Célina Ducroux

## Abstract

**BACKGROUND:** Monitoring stroke patients in critical-care units for 24 hours after thrombolysis or endovascular thrombectomy is considered standard of care but is not evidence-based. Due to the Covid-19 pandemic, our center modified its protocol in April 2021 with 24-hour critical-care monitoring no longer being guaranteed for stroke patients. We aim to compare the incidence and timing of complications over the first 24 hours post-reperfusion therapies and their association to hospital unit in 2019, 2020 and 2021.

**METHODS:** We conducted a single-center retrospective cohort study. We analyzed data from stroke patients treated with thrombolysis and/or endovascular thrombectomy at our center in 2019 (pre-Covid-19, standard of care), 2020 (during Covid-19, standard of care) and 2021 (during Covid-19, new protocol). Data extracted included demographics, the nature and timing of complications within the first 24 hours, and the unit at the time of any complication. Major complications included neurologic deterioration, symptomatic intracranial hemorrhage, recurrent stroke, myocardial infarction, systemic bleeding, rapid assessment of critical events call, and death.

**RESULTS:** Three hundred forty-nine patients were included in our study: 78 patients in 2019, 115 patients in 2020, and 156 patients in 2021. In 2021, 32% of patients experienced at least one complication within the first 24 hours compared to 34% in 2020 and 27% in 2019. In 2021, 33% of patients admitted to critical-care units had a complication compared to 31% in 2020 and 26% in 2019. In 2021, 70% of complications had occurred by hour eight compared to 49% in 2020 and 29% in 2019. Patients with low NIHSS score at presentation and treated only with tPA had significantly less complications.

**CONCLUSIONS:** Despite the change of protocol in April 2021, the incidence and timing of complications did not significantly worsen compared to prior years and were not associated with hospital location. Further research is required to evaluate the necessity of critical care monitoring for 24 hours in this population, specifically patients with low NIHSS score at presentation and treated only with tPA.

## Introduction

Since the initiation of intravenous thrombolysis with alteplase (tPA) for the treatment of acute ischemic stroke in the mid-1990s (1), it has been considered standard of care for all patients receiving thrombolysis with or without endovascular thrombectomy to be monitored in critical-care units (such as Level 1 or Level 2 intensive-care units) for 24 hours post-treatment. (2-4) This practice is resource-intensive and lacks empirical support, reflecting 30-year old concerns about lingering hypofribrinogenemia induced by tPA and the risk of intracranial hemorrhage post-thrombolysis. (5) While this risk is not negligible and is well-documented at 3-7% (6-8), the half-life of alteplase is measured on the order of minutes, (9) and it is not clear whether post-thrombolysis complications are evenly distributed among stroke patients and over the subsequent 24 hours. Similarly, there is no rationale other than tradition for 24 hours of monitoring for patients undergoing thrombectomy without thrombolysis, whose risk of hemorrhagic complications is unlikely to be present for 24 hours. (10) A handful of recent observational studies suggest that certain subgroups of patients (e.g., lower stroke severity, negative MRI post-thrombolysis) may not require admission to a critical-care unit (11-13) or may require a shorter period of intensive monitoring. (14, 15) To our knowledge, no randomized trial level data have been published supporting 24 hours of monitoring for patients post-thrombolysis or post-thrombectomy.

The third wave of the Covid-19 pandemic in early 2021 caused significant resource limitations in hospitals – especially critical care units – across Canada. (16) In response, The Ottawa Hospital modified its stroke monitoring protocol in mid-April 2021 to allow stroke patients to spend less time in critical care units or to be admitted to other units in the hospital including general neurology or internal medicine wards. In other words, stroke patients were no longer guaranteed access to 24 hours of critical-care monitoring. This pragmatic reorganization of care created the opportunity to study non-standard of care monitoring practices for acute stroke patients. This study sought to evaluate the incidence and timing of complications among stroke patients during the first 24 hours following the administration of thrombolysis and/or endovascular thrombectomy (EVT), and their association with the hospital unit to which they are admitted. Crucially, we are interested in determining if these outcomes varied significantly from past years when standard of care practices were followed. We hypothesized that, under the new stroke monitoring protocol in 2021, the incidence of major complications would be low (less than 20%), that the incidence of symptomatic intracranial hemorrhage would not exceed 7% (which is the higher-bound rate of post-thrombolysis intracranial hemorrhage in the published literature), that the majority of complications (>50%) would occur in the first 8 hours post admission, that the incidence and timing of complications would not be associated with the hospital unit to which patients are admitted, and that these findings would not vary significantly from past years.

## Methods

We conducted a single-center retrospective cohort study that analyzed clinical and imaging data gathered from a high-volume comprehensive stroke center. The study was approved by the local institutional review board. All relevant data were extracted from electronic medical records and were reviewed by two members of the research team (TLT and CD). The data supporting the findings of this study are available from the corresponding author upon reasonable request by a qualified investigator. We followed the STROBE guidelines for conducting and reporting observational research (the complete STROBE checklist can be found in Appendix A).

### Patient population

The Ottawa Hospital uses a QI infrastructure that tracks all stroke codes called to the CIVIC Campus Emergency Department (ED). This database was used to identify all patients presenting as stroke codes who received tPA and/or EVT and who were admitted to the hospital between April 15 (date when The Ottawa Hospital’s new stroke monitoring protocol was issued) and September 30 2021. Additionally, we collected the same data on patients fulfilling inclusion criteria from April 1 to September 30 2020 (first wave of COVID-19) and June 1 to September 30 2019 (pre-COVID-19). This shorter data collection period in 2019 is explained by the fact that the hospital underwent a change in electronic medical records in June 2019 that prevents our access to records before that time. This resulted in unequal sample sizes between our three years. Patients presenting as stroke codes as admitted in-patients, patients presenting as stroke codes with a diagnosis other than ischemic stroke, and patients presenting with an ischemic stroke who were not candidates for reperfusion treatment were excluded from this study. No informed consent was required as this study did not influence the care of patients.

Patient baseline characteristics (age, sex), Covid-19 status, National Institutes of Health Stroke Scale (NIHSS) score at presentation and at 24h, time of symptom onset (or last seen well), reperfusion treatment received (intravenous thrombolysis administration [tPA or tenecteplase], EVT, or both), timing of reperfusion treatment, and antiplatelets therapy received were collected.

### Outcomes

We recorded the incidence and timing of complications occurring in the first 24 hours after the stroke code was initiated. Complications were divided as major (Neurological deterioration [≥ 4 points on NIHSS], new or larger ischemic stroke on neuroimaging, hemorrhagic transformation, symptomatic intracranial hemorrhage, myocardial infarction, systemic bleeding, and death) and minor (angioedema, fever [temperature >37.9C], seizure, groin hematoma, rapid assessments of critical events (RACE) call, and move to higher unit). The incidence of hemorrhagic transformation was collected from the non-contrast head computed tomography (CT) performed for all patients 24 hours after reperfusion treatment (as per our institution’s stroke protocol) or earlier if there was a concern about symptomatic intracranial hemorrhage. Symptomatic intracranial hemorrhage was defined as any hemorrhagic transformation with neurological deterioration as indicated by a change in NIHSS ≥ 4 points compared to baseline. (17) If a patient had multiple complications, the timing of the first complication was used in the analyses. The hospital units to which patients were admitted right after reperfusion treatment, in which they were at the time of the complication, and at 24 hours were also collected. These were categorized as critical care units (medical intensive care units [ICU], Neurological Acute Care Unit [NACU], Post-anesthesia Acute Care Unit [PACU]) and non-critical care units (neurology ward, medicine ward, or emergency departments [ED-Resuscitation, ED-emergent, and ED-Observation]). Patients were considered in the non-critical care unit subgroup if they were moved to a non-critical care unit at any point during the first 24h following their stroke code initiation or if they had a complication in a non-critical care unit.

### Statistical analysis

Data were analyzed using descriptive statistics with chi-squared analysis as well as Kaplan-Meier curves with log-rank tests in accordance with the outlined questions and hypotheses. An interim analysis took place after data from the first 100 patients were collected to see if an unexpectedly high rate of complications (arbitrarily set at 40%) occurred, in which case the pandemic-related changes in practice would have been reassessed. Preplanned subgroup analysis includes analysis by treatment group (thrombolysis, EVT, and thrombolysis + EVT) and by NIHSS on presentation (under or equal to 10, and over 10).

## Results

### Demographics

Three hundred forty-nine patients were included in our study: 78 patients in 2019, 115 patients in 2020 and 156 patients in 2021. Baseline characteristics of the patients are shown in Table 1. Patient age, sex, median NIHSS, door-to-needle times and door-to-groin puncture times showed no significant differences. In 2021, 137 patients (87.8%) were hospitalized in critical care units compared to 106 patients (92.2%) in 2020 and 56 patients (71.8%) in 2019.

**Table 1:** Baseline characteristics of patients.

| Characteristics | 2019<br>N=78 | 2020<br>N=115 | 2021<br>N=156 | p-value |
| --- | --- | --- | --- | --- |
| Age (mean±SD) | 72.8±12.1 | 69.5±15.5 | 72.6±13.9 | 0.14 |
| Female n (%) | 37 (47.4) | 42 (36.5) | 67 (42.9) | .30 |
| Covid screening n (%) | - | 82 (71.3)<br>(n=82) | 124 (79.5)<br>(n=124) | - |
| Negative | - | 69 (60.0) | 94 (75.8) | .15 |
| Suspect | - | 13 (11.3) | 29 (23.4) | - |
| Positive | - | 0 | 1 (0.8) | - |
| **Stroke event** |  |  |  |  |
| Baseline NIHSS; median [IQR] | 13 [8 - 19] | 12 [7.75 - 18.0]<br>(n=114) | 14 [7 - 18.5]<br>(n=153) | .52 |
| Thrombolysis n (%) | 62 (79.5) | 84 (73.0) | 99 (63.5) | - |
| tPA | 62 (79.5) | 84 (73.0) | 79 (50.6) | - |
| TNK | - | - | 20 (20.2) | - |
| EVT n (%) | 42 (53.8) | 69 (60) | 90 (57.7) | - |
| Both Thrombolysis and EVT | 26 (33.3) | 38 (33.0) | 33 (21.1) | - |
| Door-to-needle; min; median [IQR] | 35.0 [28.4 - 50.7]<br>(n=72) | 34.0 [27.7 - 47.2]<br>(n=108) | 31.1 [25.5 - 44.5]<br>(n=145) | 0.32 |
| Door-to-groin puncture, min; median [IQR] | 72.4 [41.2 - 103.5] | 65.1 [48.1 - 80.7] | 53.0 [36.7 - 92.1]<br>(n=150) | 0.43 |
| Antiplatelet n (%) | 24 (30.8) | 19 (16.5) | 31 (19.9) | - |
| None | 54 (69.2) | 96 (83.5) | 125 (80.1) | - |
| ASA | 12 (15.4) | 6 (5.2) | 19 (12.2) | - |
| Clopidogrel | 1 (1.3) | 1 (0.8) | 0 (0) | - |
| DAPT | 7 (9.0) | 6 (5.2) | 10 (6.4) | - |
| Integrillin | 4 (5.1) | 6 (5.2) | 2 (1.3) | - |
| **Hospitalization Unit** |  |  |  |  |
| **After reperfusion treatment** |  |  |  |  |
| ICU n (%) | 56 (71.8) | 106 (92.2) | 137 (87.8) | - |
| non-ICU n (%) | 22 (28.2) | 9 (7.8) | 19 (12.2) | - |
| **At 24 hours** |  |  |  |  |
| ICU n (%) | 59 (75.6) | 111 (96.5) | 134 (85.9) | - |
| non-ICU n (%) | 19 (24.4) | 4 (3.5) | 22 (14.1) | - |

### Complications

The rate of complications after 100 patients in 2021 was 37%, which did not meet our predetermined threshold for reassessing the modified protocol. Table 2 summarizes the main findings of the study. In 2021, 50 (32%) patients had, at least, one complication compared to 39 (34%) in 2020 and 21 (27%) in 2019, p=0.58. Thirty-seven patients (24%) had a major complication in 2021, compared to 36 patients (31%) in 2020 and 17 patients (22%) in 2019, p=0.25. Rates of hemorrhagic transformation were not significantly different across the years: in 2021, 24 patients (15%) had a hemorrhagic transformation compared to 26 patients (23%) in 2020 and 11 patients (14%) in 2019, p=014. Only the difference in incidence of symptomatic intracranial hemorrhage across the three years was statistically significant: in 2019, there were six symptomatic intracranial hemorrhage (8%) compared to two (2%) in 2020 and three in 2021 (2%), p=0.03.

**Table 2:**
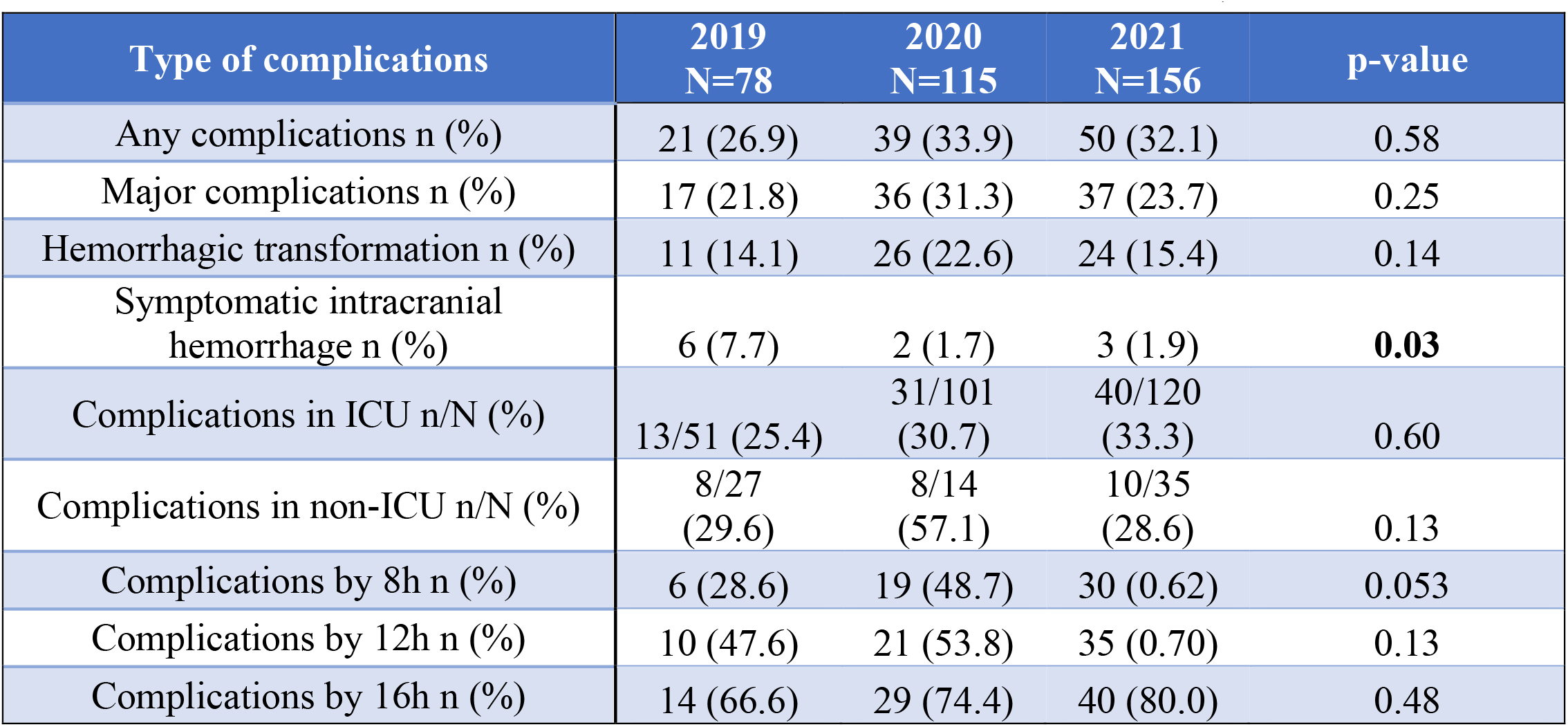
Complications after reperfusion therapy within 24 hours in 2019, 2020 and 2021.

### Complications by Time

Figure 1 represents the timing of complication over the first 24 hours of care for all three years as well as based on the unit to which patients were admitted. Across all three years, 60% of complications occurred within 12 hours. Across all three years, 29% of complications occurred in the first 8 hours in 2019 compared to 49% in 2020 and 60% in 2021, *p* = .053, which trended towards statistical significance. This trend disappeared with time. By twelve hours, 48% of complications had occurred in 2019 compared to 54% in 2020 and 70% in 2021, *p* = .13. By sixteen hours, 67% of complications had occurred in 2019 compared to 74% in 2020 and 80% in 2021, *p* = .48. Otherwise, 46% of symptomatic intracranial hemorrhage occurred within 16h across all three years. In 2021, all symptomatic intracranial hemorrhage occurred within 12h. In 2020, none of the two symptomatic intracranial hemorrhage occurred within 12h. In 2019, 33% of symptomatic intracranial hemorrhage occurred within 12h.

**Figure 1.**
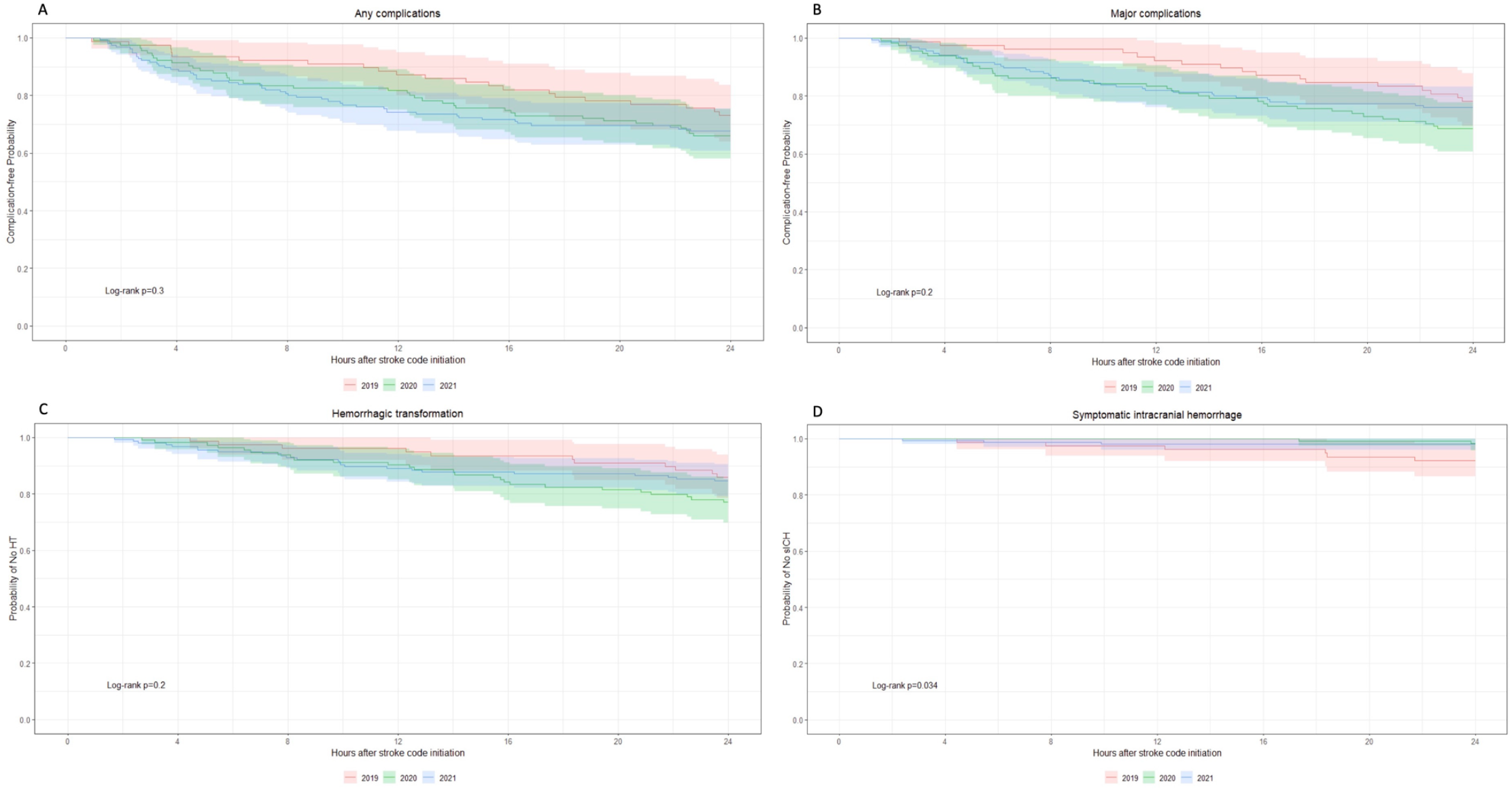
Rate of complications in the first 24 hours following stroke code initiation in acute stroke patients treated with reperfusion therapies between 2019, 2020 and 2021.

### Complications by Unit

Forty of the complications (80%) in 2021 occurred in patients admitted to critical care units compared to 36 (80%) in 2020 and 16 (62%) in 2019. The incidence of all complications that occurred in patients admitted to critical care units was 33% in 2021, 31% in 2020 and 25% in 2019, *p* = .60. The incidence of all complications that occurred in non-critical care units was 29% in 2021, 57% in 2020 and 30% in 2019, *p* = .13. In ICUs, 62% of complications occurred within 12h. In non-critical care unit, 73% of complications occurred within 12h. There was a statistical association in 2020 between the unit of admission and the incidence of any complication (log-rank p = 0.01) and major complications (log-rank p = 0.05). However, this association was not present in 2021 or 2019 (Figure 2). There was no statistical difference in the rate of complications between all three years in critical care units (log-rank p = 0.2), but a trend towards statistical significance in non-critical care units with a higher rate of complications in 2020 compared to 2021 and 2019, log-rank p=0.08 (Figure 3).

**Figure 2.**
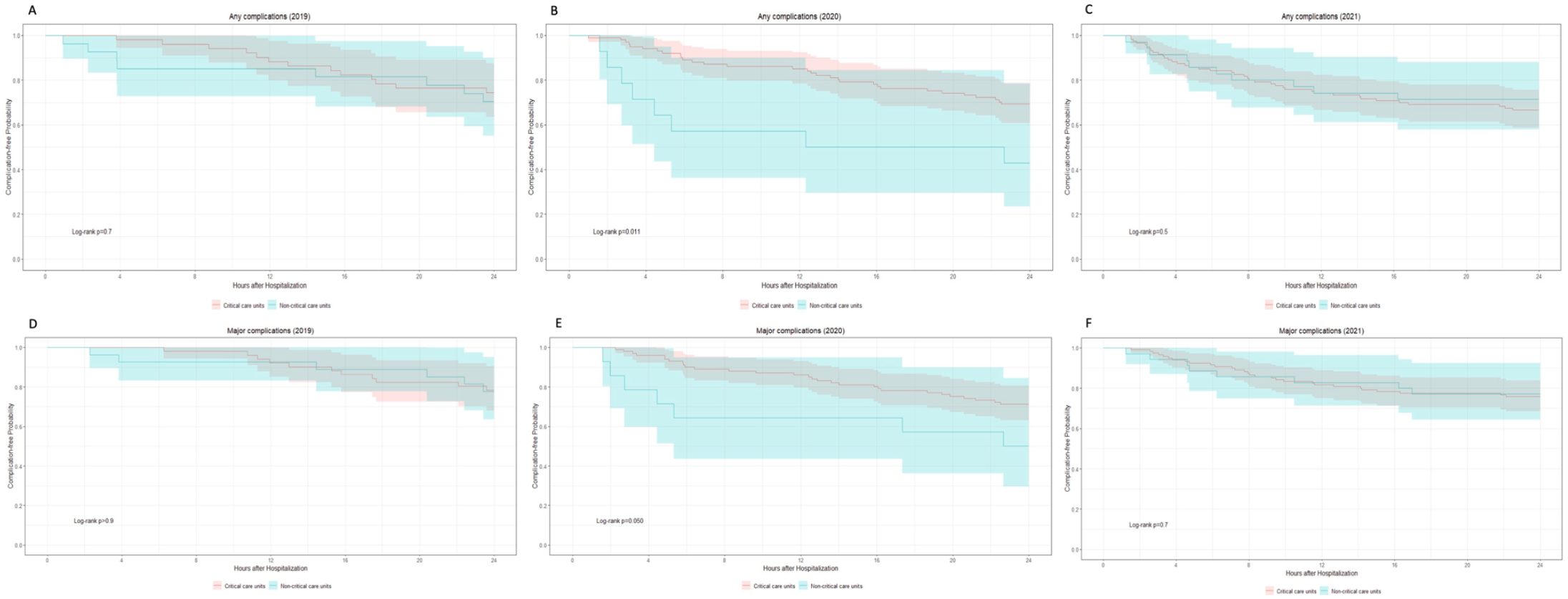
Rate of complications over the first 24 hours following stroke code initiation in acute stroke patients based on unit of admission in 2019, 2020 and 2021.

**Figure 3.**
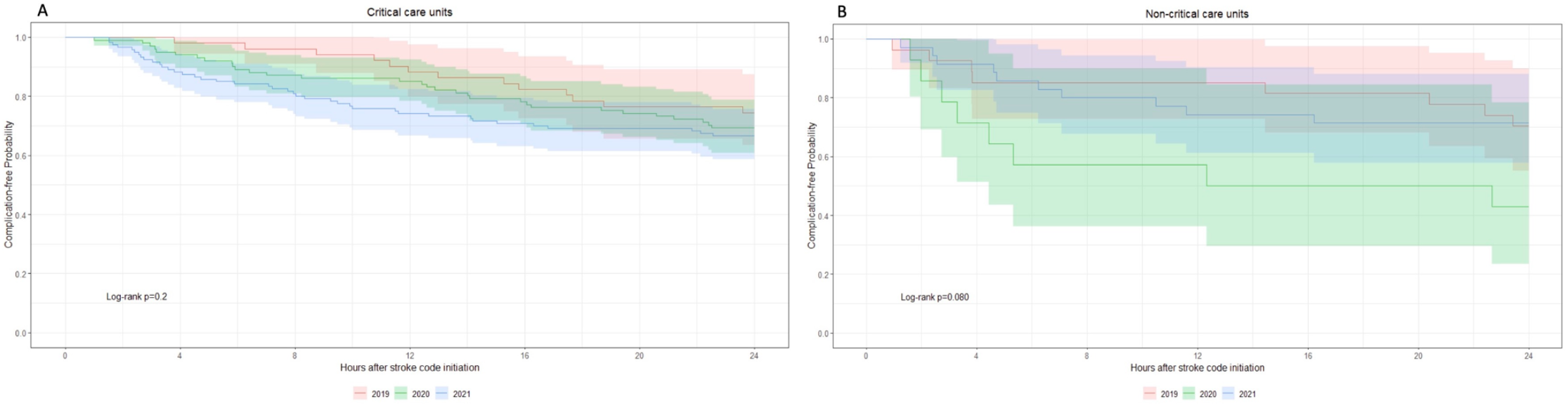
Rate of any complications over the first 24 hours following stroke code initiation in acute stroke patients based on unit of admission in 2019, 2020 and 2021.

### Complications by treatment groups

The incidence of any complications, of major complications, and of hemorrhagic transformation was lower in the thrombolysis group while the incidence of symptomatic intracranial hemorrhage was lower in the EVT group (Figure S1-S2). Over all three years, the incidence of any complication was 23% in the thrombolysis group, 34% in the EVT group and 41% in the combined therapy, log-rank p=0.01. The incidence of major complications followed a similar pattern with 20% for the thrombolysis group, 28% for the EVT group and 32% for the combined therapy, but the difference was not significant, log-rank p=0.08. The incidence of hemorrhagic transformation was lower in the thrombolysis group at 11.0% compared to the EVT group (21%) and the combined therapy (24%), log-rank p=0.04. Lastly, the incidence of symptomatic intracranial hemorrhage was not significantly different between the groups with 3% in the EVT group, 4% in the thrombolysis group, and 3% in the combined therapy group, log-rank p>0.9.

### Complications by NIHSS

When patients were analyzed by dichotomized NIHSS (above and below 10), several statistically significant differences were identified (see Figure S3-S4). Of note, the median NIHSS for the patients with NIHSS ≤ 10 was 7, compared to 18 for the group of patients with NIHSS > 10. The incidence of any complication was 22% for patients presenting with a NIHSS ≤ 10 compared to 38% for NIHSS > 10, log-rank p=0.002. The incidence of major complications was 17% for NIHSS ≤ 10 and 31% for NIHSS > 10, log-rank p=0.006. The incidence of any intracranial hemorrhage was 10% for NIHSS ≤ 10 and 22% for NIHSS > 10, log-rank p=0.013. Lastly, the incidence of symptomatic intracranial hemorrhage was higher for NIHSS > 10 with 4% compared to 2% for NIHSS ≤ 10, but this difference was not significant, log-rank p=0.15. In the low NIHSS group, 66% of complications had occurred by hour 8 compared to 44% in the high NIHSS group, p=0.05.

## Discussion

This study took advantage of a natural variation in clinical care brought about by COVID-19 protocols at our center to assess the incidence and timing of complications among stroke patients receiving thrombolysis and/or EVT in relation to the hospital unit to which they were admitted. As such, this study provides the opportunity to interrogate the standard of care according to which stroke patients undergoing reperfusion therapies should receive 24 hours of critical-care monitoring.

Our study specifically investigated how complications varied based on unit of admission in the context of a pandemic-driven protocol change in 2021. Had the quality of care in 2021 been inferior due to the admission of post-thrombolysis and post-thrombectomy patients to non-critical care units, this may have manifested with a higher incidence of complications due to substandard care, or a falsely lower incidence due to lack of identification, or identified later. Ultimately, we saw no significant difference either in incidence or severity of complications. The incidence of any complications and major complications was similar across all three years. The only notable difference was a higher rate of symptomatic intracranial hemorrhage in 2019 (7.7%), prior to any change in protocol. There was also no difference in the timing of complications: if anything, complications were identified earlier in 2021 than in previous years. For instance, by 8 hours after the stroke code,62% of complications had occurred in 2021 compared to 49% in 2020 and 29% in 2019, which trended towards significance (p=0.053).

Finally, no association was found between the incidence of complications and the unit to which patients were admitted in 2021.These results suggest that 24 hours of critical care monitoring may not be necessary in all cases of stroke post thrombolysis and thrombectomy. At the very least, these results could support and accelerate the development of low-intensity monitoring protocols for subgroups of stroke patients following reperfusion treatment such as the one being developed by Roland Faigle at John Hopkins University. (18, 19)

Overall, one-third (32%) of the patients included in our study suffered at least one complication in the first 24 hours following reperfusion treatment. This was higher than hypothesized, though no other studies were found reporting the incidence of all complications at 24 hours for this population. However, the incidence of symptomatic intracranial hemorrhage in our study at 3% is within the documented risk of 3-7% in the literature. (6-8) Patients with a NIHSS ≤ 10 were generally half as likely to suffer a complication compared to patient with a NIHSS > 10. Similarly, the rate of symptomatic intracranial hemorrhage jumped from 1% for NIHSS≤ 10 to 4% for NIHSS > 10. This is similar to other studies that support the proposition that patients with higher NIHSS are more likely to experience neurological deterioration or to require ICU resources. (13, 15) The incidence of any complication was highest in the combined therapy groups and lowest in the thrombolysis only group. The incidence of symptomatic intracranial hemorrhage was similar in the thrombolysis and combined therapy groups, and are comparable to the findings of the major trials comparing EVT to EVT and thrombolysis. (20, 21) However, the incidence of symptomatic intracranial hemorrhage was lower in the EVT group (2%) compared to the other two groups (3-5%). This differs from the published literature showing that the rate of symptomatic intracranial hemorrhage is not different when EVT is used alone or in combination to thrombolysis. (22, 23) Recent observational studies have investigated the timing of complications following thrombolysis and showed that over 80% of symptomatic intracranial hemorrhage and neurological deterioration occurs before 12 hours post-treatment. (14, 15) Our study showed similar findings in 2021 with 80% of any complications and all three symptomatic intracranial hemorrhage occurring within 12 hours post-treatment. A randomized controlled trial (RCT) led by Dr. Craig Anderson in Australia is exploring the question of 8 hours of post-tPA monitoring versus the standard 24 hour monitoring but is currently only at the recruitment phase. (24)

We acknowledge several limitations to our study. First, the sample sizes across the three years are unequal, as explained in our methods section. Second, it is important to note that we cannot draw any causal connections between incidence of timing of complications and the unit of admission because of the selection bias present in our study. This study was observational by design and as such patients were not randomly assigned to critical care or non-critical care. In 2019 and 2020 when standard of care was followed, patients would normally be guaranteed critical care monitoring unless their beds were not available. In 2021, because of the protocol change, patients were no longer guaranteed critical care monitoring, but the stroke neurologist on duty still had to make an informed decision as to where they believed the patient would best be cared for. While the rationale behind the allocation of patients to critical care units differed in 2021, the actual number of patients hospitalized in critical care units in 2021 (80%) was comparable to 2020 (79%) and higher than 2019 (62%). This limits the interpretation of our results. Third, the single-center nature of the study does limit the generalizability of our results. Finally, the modified Rankin Score (mRS) at three months was not collected for most patients, which limits the interpretation of the long-term clinical impact of our study. For instance, even if complications were potentially detected earlier in 2021 compared to prior years, it is unclear whether this led to a change in long-term outcomes.

In conclusion, the change of protocol in April 2021 does not appear to have impacted negatively the care of stroke patients at our center in the 24 hours after thrombolysis or thrombectomy: the overall incidence of complications did not increase compared to previous years, there was no association between the incidence of complications and the unit of admission in 2021, and complications were not detected later in 2021 compared to previous years.

Monitoring stroke patients for 24 hours in critical care requires significant investment of resources and may not be required in all cases. Subgroups of patients - particularly those with lower NIHSS on presentation or receiving thrombolysis only - could be monitored in critical care for shorter periods of time or be monitored in lower intensity units. Other predictors that could be investigated include age, comorbidities, and time of symptom onset. We hope this study will contribute to a broader discussion and motivate further research on the necessity of 24 hours of critical care level monitoring for this population.

## Data Availability

The data supporting the findings of this study are available from the corresponding author upon reasonable request by a qualified investigator.

## Non-standard Abbreviations and Acronyms

tPA: altepase
EVT: endovascular therapy
NIHSS: National Institutes of Health Stroke Scale
ED: emergency department
RACE: rapid assessment of critical events
CT: computed tomography
ICU: intensive care units
NACU: Neurological Acute Care Unit
PACU: Post- anesthesia Acute Care Unit.

## Acknowledgements

I would like to thank co-authors Dr. Ducroux and Dr. Shamy for their ongoing support as well as Dr. Ramsay and his team for their help with the statistical analysis.

## Supplemental Material

Figure S1-S4

